# The transition to retirement and dementia risk: An application of sequence analysis

**DOI:** 10.64898/2026.10.07.26364998

**Authors:** Lucia Pacca, Jillian Hebert, Dawn C. Carr, Willa D. Brenowitz, M. Maria Glymour, Amal Harrati, Anusha M. Vable

## Abstract

**Background:** Retirement is a major life transition that may influence cognitive aging. Retirement pathways are becoming increasingly diversified, yet evidence on how heterogeneous retirement trajectories relate to dementia risk remains limited. This study used sequence and cluster analysis to characterize thousands of transition-to-retirement trajectories, including retirement timing and work intensity, to estimate associations with dementia risk.

**Methods:** We used data from 13,879 participants in the U.S. Health and Retirement Study. Our exposure was transition-to-retirement trajectories between ages 51 and 75 identified using sequence and cluster analysis. Our outcome was a validated algorithmically derived dementia probability measure. We estimated associations between trajectory clusters and dementia risk beginning at age 76 using generalized estimating equations, adjusting for sociodemographic characteristics. Inverse probability weights were used to account for attrition.

**Results:** We identified ten distinct transition-to-retirement trajectories characterized by differences in timing of retirement, part-time work, disability, and labor force attachment. Compared with individuals retiring after full-time work in their early-to-mid 60s, dementia risk was similar among those retiring earlier or later from full-time work. In contrast, individuals with sustained part-time employment between ages 51 and 75 had lower odds of dementia (OR: 0.62, 95% CI: 0.48, 0.80), while trajectories characterized by prolonged disability were associated with higher odds of dementia (OR: 1.53, 95% CI: 1.22, 1.91).

**Conclusions:** Retirement timing alone showed limited association with later-life dementia risk, whereas sustained part-time employment and disability-related trajectories were more strongly associated with dementia risk. These findings suggest that employment intensity and health-related exit from the labor force may be more relevant for dementia risk than retirement timing, and that gradual and flexible work-to-retirement transitions may benefit brain health.

## Background

The number of new dementia cases in the United States is projected to nearly double between 2020 and 2060 as the population ages^1^. Given the lack of effective treatment for dementia, identifying modifiable factors associated with dementia risk is an important prevention priority^2^. Employment-related and social factors have been associated with dementia risk^2–4^ and may be potentially relevant for prevention.

Due to longer life expectancy, the expected years of life after retirement has increased worldwide. For men in the United States, the length of life after retirement has increased from 12.8 years in 1970 to 18.6 years in 2023. For women, retirement length increased a similar amount, from 16.6 years in 1970 to 21.3 years in 2023^5^. Many countries, including the United States, are adapting to increasing longevity by gradually raising retirement age. Some theories suggest that timing of retirement may affect cognitive functioning. According to the “use it or lose it”^6^ and “mental retirement” hypotheses,^7^ continued engagement in cognitively demanding activities helps preserve cognitive functioning, whereas reduced cognitive stimulation may contribute to cognitive decline. Paid employment may provide an important source of such stimulation through complex tasks, problem solving, learning, and other cognitively demanding activities. Retirement may reduce opportunities for cognitive engagement previously provided by work, with earlier retirement potentially extending the period of reduced work-related cognitive stimulation. However, existing findings on the association between retirement timing and cognition are inconclusive. Several prior studies found a negative effect of retirement on cognition^7–9^, but other studies found no association between retirement age and cognition^10,11^ or even that later retirement was related to worse cognitive performance^12^. One potential explanation of prior inconsistent findings is that conventional measures of retirement timing obscure substantial heterogeneity in how individuals transition out of paid work. Two individuals retiring at the same age may reach retirement through very different pathways, with different durations and timing of full-time work, part-time work, unemployment, disability, and labor force re-entry, and these differences may be relevant for cognitive aging. Interpretation of prior findings may be further complicated by self-selection. Observed associations between earlier retirement and cognitive outcomes may partially reflect health-related selection into retirement, as poor health or disability can be reasons for earlier retirement and are themselves associated with cognitive decline^13,14^.

In addition to timing differences, retirement pathways are becoming increasingly diversified^15^. Partially influenced by growing economic insecurity and wealth inequality, increasing numbers of older individuals experience more gradual exits from the labor force or return to paid work, often part-time, after retirement^16^. Part-time work during the transition-to-retirement years may be sufficient to maintain cognitive stimulation while at the same time facilitating engagement in other activities outside of work, which may be beneficial for cognition. Compared to full-time work, part-time work may be associated with lower exposure to stress, a factor that can negatively impact cognition through allostatic load^17^. However, part-time workers often have lower wages, fewer benefits, and higher financial insecurity, all factors linked to poorer cognitive health. A recent study on an older cohort of Americans found that, while full retirement was associated with worse cognitive outcomes compared to full-time employment, partial retirement was associated with similar outcomes to full-time employment^18^. However, very limited literature examines longitudinal trajectories incorporating both retirement timing and work intensity (part-time vs. full-time work) during the transition-to-retirement years, obscuring potentially differential relationships between retirement paths and cognitive aging.

Prior studies have either evaluated timing of retirement (by evaluating retirement age) or classified different retirement paths based on the type of transition (e.g., full retirement vs. partial retirement) but without fully incorporating the complexity of patterns of labor market transitions over several years. Although a small number of studies have used trajectory-based approaches to characterize longer-term retirement pathways, these studies have generally focused on the determinants or socio-economic consequences of retirement trajectories rather than their associations with cognitition^19–21^. We fill several key gaps here. First, we characterize transition-to-retirement patterns incorporating timing of retirement as well as differences in timing, duration and order of different levels of work intensity (part-time vs. full-time work). We then evaluate whether those patterns are associated with later-life dementia risk. In particular, we test: [1] whether retirement timing is differentially associated with dementia risk; and [2] whether reduced work intensity during the retirement transition is associated with lower or higher dementia risk. A lower risk would be consistent with the hypothesis that reduced work intensity preserves some of the cognitive stimulation associated with employment while allowing greater engagement in activities outside of work that may benefit cognition, whereas a higher risk could reflect socioeconomic disadvantage associated with retirement pathways, including constrained opportunities for full-time work or financial constraints on retirement. We use data from the Health and Retirement Study (HRS) and identify clusters with similar transition-to-retirement patterns between ages 51-75 using sequence and cluster analysis. We then estimate the associations between transition-to-retirement clusters and dementia risk at age 76 and older. By capturing heterogeneities in transition-to-retirement trajectories, this novel application of sequence analysis provides insight into how the timing and intensity of labor force exit shape later-life dementia risk, with implications for policies supporting healthy cognitive aging.

## Methods

### Sample

Our sample included respondents in the Health and Retirement Study (HRS), a nationally representative longitudinal study of U.S. adults aged 50 and older and their spouses of any age. The HRS began in 1992, with participants interviewed approximately every two years and new cohorts added periodically to maintain population representativeness. Across the 15 waves included in this analysis (1992–2020), 43,558 participants entered the HRS. We restricted the eligible sample to 16,949 participants who survived and were in the HRS sample at age 76. Of these, 13,966 had at least one dementia probability score available at age 76 or older during the outcome assessment period (2000–2020). We selected age 76 as the beginning of the outcome assessment period to allow employment and retirement trajectories to be characterized through age 75, capturing the transition to retirement for most individuals before assessment of dementia outcomes. These 13,966 participants comprised the sequence-analysis sample. For analyses examining associations between retirement trajectories and later-life dementia risk, we excluded 87 participants with missing covariate information, resulting in a final analytic sample of 13,879 participants (49,360 person-waves; Appendix Figure 1).

### Exposure Measure

Our exposure was transition-to-retirement clusters obtained using sequence and cluster analysis from self-reported employment information. Employment status was reported at each wave. Retrospectively, participants also reported the year they began their current job and retired, as well as the start and end years of up to three previous jobs lasting at least five years. The sequence and cluster analysis consisted of three steps.

### 1 Creating and cleaning individual transition-to retirement sequences

We constructed annual employment sequences spanning ages 51–75 for all respondents, regardless of their age at study entry, using retrospective employment information to characterize years preceding HRS enrollment when available. Each year from ages 51-75 was characterized by one of 7 mutually exclusive categories (or “states”): full-time work, part-time work, retirement, disability, not working or unreported. The not working category included those who were taking care of family members (7.8%), unemployed (2.3%), or on temporary leave from work (1.0%). These three categories were merged due to small percentages of time points spent in unemployment or on temporary leave. Similar to prior work^21,22^, we used self-reported, prospective information on employment status reported at each survey wave as the main variable to determine states. When prospective information was not available, including for years preceding a respondent’s entry into the HRS, we imputed states as follows. First, we imputed missing states due to off-survey years by carrying forward or backward adjacent values when the preceding and subsequent employment states were identical. Second, we used retrospective information on prior jobs and retirement, when available, to impute the remaining missing states. States that could not be reconstructed from these sources - most commonly for periods before respondents entered the HRS when prior employment histories were unavailable - were categorized as”unreported” (19.2% of states).

### 2 Sequence analysis to quantify differences between trajectories

We calculated the dissimilarity between individual trajectories using the Optimal Matching algorithm^23^. This algorithm computes the dissimilarity between each pair of trajectories as the combination of substitution, insertion or deletion “costs,” corresponding to edit operations to make two sequences equal. Substitution costs between states were based on transition frequencies, such that states with more frequent transitions between consecutive time points were assigned a lower cost. Insertion and deletion costs were set to 1, equivalent to half the maximum substitution cost. The result of the sequence analysis was a dissimilarity matrix comparing each trajectory to all the other trajectories in the data with larger numbers representing more dissimilar trajectories

### 3 Cluster analysis to group similar trajectories

We conducted cluster analysis on the dissimilarity matrix to group similar transition-to-retirement trajectories using hierarchical clustering. We selected the optimal number of clusters based on cluster quality indicators, which allowed selecting clusters that were more homogeneous within themselves and more distinct from other clusters

### Outcome Measure

We used an algorithmically defined dementia probability score (ranging from 0-1) developed by Gianattasio et al. based on the Aging, Demographics, and Memory Study (ADAMS)^24^. This measure, available for HRS participants 70 and older, harmonizes data from direct respondents and proxy respondents to the HRS surveys, therefore retaining the most cognitively impaired individuals, who would typically be excluded due to missing data. Dementia probability scores were predicted using demographics (age, sex and race) and cognitive assessments (immediate and delayed word list recall, Telephone Interview for Cognitive Status and the Jorm Informant Questionnaire for Cognitive Decline). The Gianattasio measure offers several advantages, including the incorporation of proxy respondents to reduce selection bias, explicit development to achieve comparable sensitivity and specificity across race and ethnicity groups, extended coverage of the HRS through 2020, and a continuous probability scale that captures gradations in cognitive impairment^24^. We included repeated measurements of dementia probability starting at age 76, one year after the end of the exposure window.

### Covariates

We selected a set of variables that preceded the transition-to-retirement trajectories and are likely to influence both retirement trajectories and dementia risk. These included gender (male or female), age, birthplace (US non-South, Southern US, foreign birth, or US non-specified), race and ethnicity (non-Hispanic white, non-Hispanic black, Hispanic), years of schooling (5-17) and mother’s and father’s education (operationalized as years of schooling). For the AHEAD cohort (participants born between 1890 and 1923), whose mother’s and father’s education were dichotomized, we used imputed values from Vable et al..^25^ We imputed missing parental education as the mean and included two separate missing indicators on mother’s and father’s education. Missing parental education may be indicative of lower childhood social capital (e.g., parents not present in the household). Although retrospective employment information allowed us to reconstruct employment states before HRS entry, some respondents entered the HRS after age 51. Therefore, we did not include covariates measured at age 51 because these were not available for all participants.

### Statistical Analysis

We estimated the association between transition-to-retirement clusters and dementia probability using generalized estimating equations (GEE) to account for repeated dementia probability scores for individuals across survey waves, with a logit link and an independent correlation structure. To account for dropout during the outcome measurement period while treating death as truncation, we used regression conditioning on continuation with inverse-probability-weighted generalized estimating equations implemented using *xtrccipw* in Stata^26^. At each time point, the probability of remaining under observation was estimated conditional on prior dementia probability and baseline predictors of attrition, including retirement-trajectory group, sex, race and ethnicity, educational attainment, mother and father’s education, missing mother and father’s education, and childhood health. The resulting inverse probability weights were incorporated into the longitudinal outcome model. We adjusted for all the covariates listed above, in addition to age of cognitive assessment and survey wave, which allows accounting for trends in dementia prevalence.

### Sensitivity Analyses

We conducted one sensitivity analysis on the exposure and one on the outcome. On the exposure, we applied an alternative algorithm to conduct sequence analysis. There are multiple methods to measure dissimilarity between trajectories, each of which has different sensitivity to different dimensions (timing, order or duration of events). The choice is usually made based on which method seems more appropriate for the research question and on the interpretability of the final clusters^22^. To assess the robustness of our analysis to a different dissimilarity measure, we conducted a sensitivity analysis using the Hamming distance^27^, which has stronger sensitivity to timing compared to duration and order of events^28^. This approach uses only substitution costs, which were set to be inversely proportional to the transition rates between states at each time point.

On the outcome, we used the Hudomiet et al. dementia probability measure, which, unlike the cross-sectional Gianattasio algorithm, is based on a longitudinal latent-variable model that incorporates repeated cognitive assessments over time, providing a complementary approach to measuring longitudinal cognitive functioning.

## Results

### Transition-to-retirement clusters

We identified 10 transition-to-retirement clusters (Figure 1) based on cluster quality measures (Figure A1). The first three clusters were mostly characterized by full-time employment and retirement, with differences in timing of retirement: 1) Retirement after full-time work at ages 62-65 (N=3,783), the typical retirement age range in the United States; 2) Retirement after full-time work before age 60 (N=2,841); and 3) Retirement after full-time work at age 70 or older (N=1,798). Clusters 4-6 were characterized by periods of part-time work, with differences in timing and duration of part-time work and retirement: 4) Gradually retired at age 70 or older (N=595), including those who had a gradual transition from full-time work to retirement, with 5-10 years of part-time work in between; 5) Retired after part-time work at age 62-65 (N=548); and 6) Continuous part-time work (N=518), characterized by either part-time employment until age 75, or retirement at age 70 or older after part-time work. Cluster 7, Mostly not working (N=396), included participants who were not working for long periods of time, mostly for family caregiving reasons. Cluster 8, Mostly disability (N=757), was characterized by periods of disability, often followed by retirement after age 65. Finally, clusters 9 and 10 were characterized by periods of unreported employment information: 9) Unreported until age 62, then retired (N=1,419); and 14) Mostly unreported (N=1,224).

**Figure 1.**
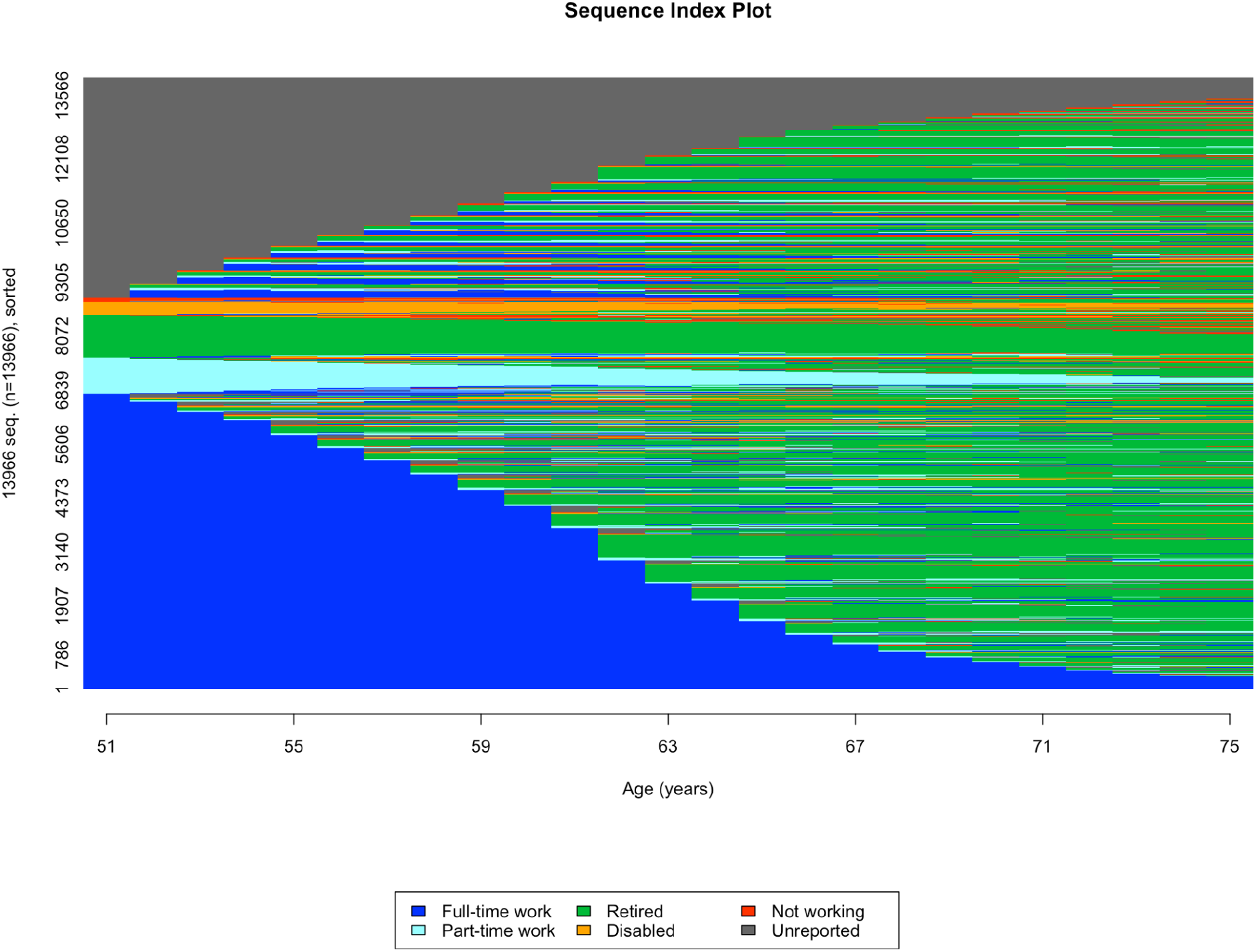
Sequence index plot for the overall sample. Individual-level transition-to-retirement trajectories built from the Health and Retirement Study. Each row on the y-axis represents one individual; each time point on the x axis is one year between age 51–75. Our sample included 13,966 individuals. Each year between age 51–75 was characterized as one of the following “states” (or categories): employed full-time, employed part-time, retired, disabled, not working or unreported.

**Figure 2.**
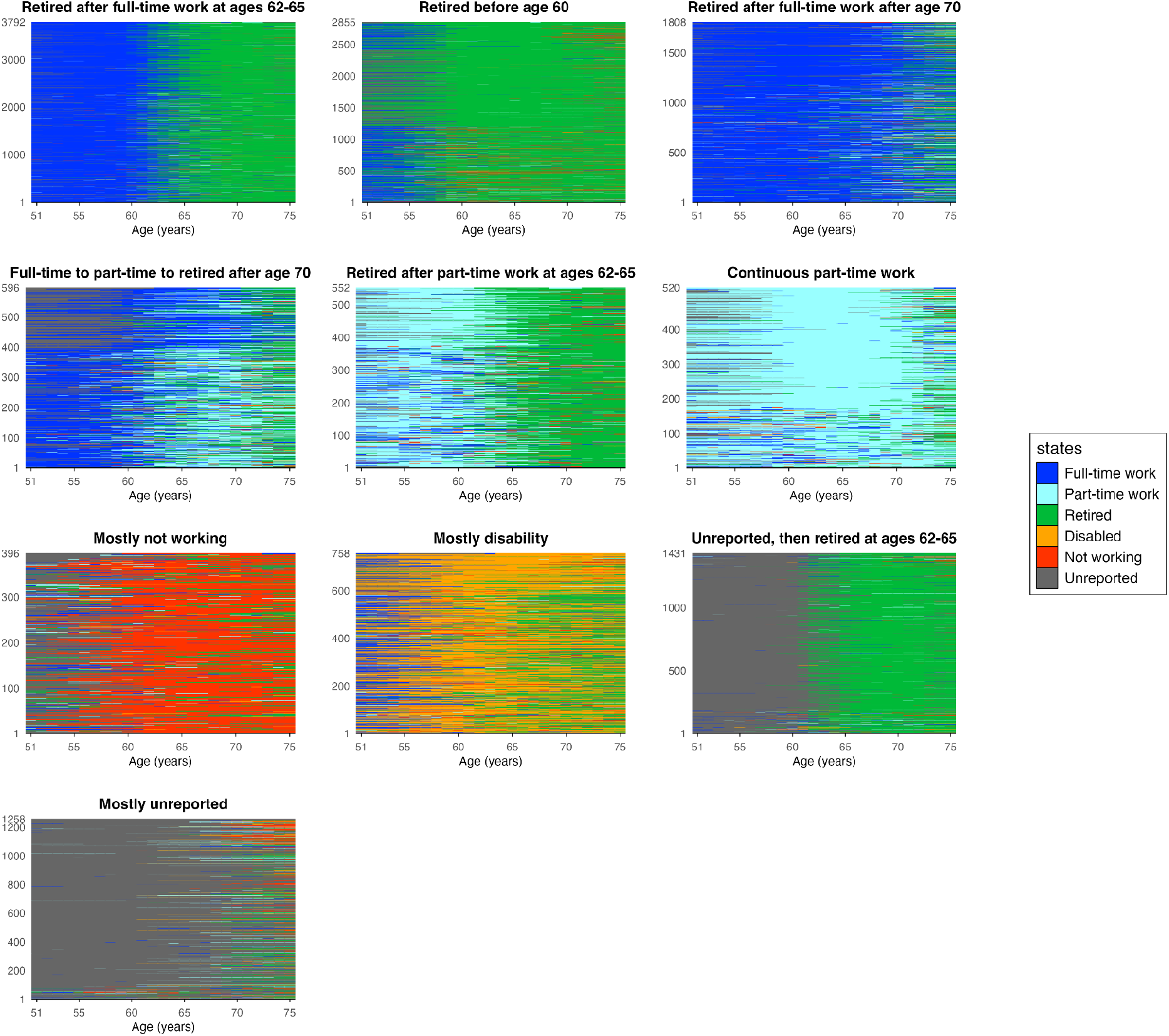
Sequence index plots by cluster. Sequence index plots for the transition-to-retirement clusters obtained using sequence and cluster analysis among HRS participants who had at least one valid observation of dementia probability score at age 76 or older (N=13,966). We applied the Optimal Matching algorithm to calculate the dissimilarity between sequences. We then implemented hierarchical clustering to group sequences based on their similarity. Based on cluster quality indicators, we identified 10 transition-to-retirement clusters. Each index plot shows the number of participants on the y-axis and age (51-75) on the x-axis.

### Summary statistics

Table 1 presents participant characteristics by transition-to-retirement cluster. Compared with the “Retired after full-time work at ages 62–65” cluster (44.8% female), most clusters had a higher proportion of female participants, with the exception of the “Retired after full-time work after age 70” cluster (41.8%). The “Mostly not working” cluster had the highest proportion of female participants (97.5%) and Hispanic participants (25.5%). The “Mostly disability” cluster had the highest proportion of non-Hispanic Black participants (29.6%) and participants born in the southern U.S. (54.1%), whereas non-Hispanic White participants were most represented in the “Continuous part-time work” cluster (83.1%). Education was generally lower among participants in the “Mostly not working,” “Mostly disability,” and clusters characterized by unreported employment information. The “Mostly disability” cluster had the lowest mean participant education (10.2 years) and, along with the “Mostly not working” cluster, among the lowest levels of parental education. Childhood health was generally similar across clusters characterized by retirement from full- or part-time work, but was poorer among the “Mostly disability” cluster and, to a lesser extent, the “Mostly not working” and clusters characterized by unreported employment information.

**Table 1.** Summary statistics by cluster.

| Characteristic | 1 | 2 | 3 | 4 | 5 | 6 | 7 | 8 | 9 | 10 |
| --- | --- | --- | --- | --- | --- | --- | --- | --- | --- | --- |
| **No. of participants** | 3,792 | 2,855 | 1,808 | 596 | 552 | 520 | 396 | 758 | 1,431 | 1,258 |
| **Age, years, mean (SD)** | 76.6 (0.7) | 76.9 (1.8) | 76.7 (1.1) | 76.7 (1.3) | 76.5 (1.0) | 76.7 (1.3) | 76.5 (0.5) | 76.5 (0.7) | 77.9 (2.8) | 78.2 (3.4) |
| **Female, No. (%)** | 1,698 (44.8) | 1,919 (67.2) | 755 (41.8) | 324 (54.4) | 412 (74.6) | 337 (64.8) | 386 (97.5) | 474 (62.5) | 846 (59.1) | 1,024 (81.4) |
| **Race/ethnicity** |  |  |  |  |  |  |  |  |  |  |
| **NH White** | 3,022 (79.7) | 2,363 (82.8) | 1,450 (80.2) | 476 (79.9) | 439 (79.5) | 432 (83.1) | 274 (69.2) | 412 (54.4) | 1,088 (76.0) | 968 (76.9) |
| **NH Black** | 520 (13.7) | 329 (11.5) | 231 (12.8) | 81 (13.6) | 80 (14.5) | 70 (13.5) | 21 (5.3) | 224 (29.6) | 187 (13.1) | 149 (11.8) |
| **Hispanic** | 250 (6.6) | 163 (5.7) | 127 (7.0) | 39 (6.5) | 33 (6.0) | 18 (3.5) | 101 (25.5) | 122 (16.1) | 156 (10.9) | 141 (11.2) |
| **Birth Place** |  |  |  |  |  |  |  |  |  |  |
| **U.S., not South** | 2,535 (66.9) | 1,908 (66.9) | 1,145 (63.5) | 405 (68.1) | 399 (72.4) | 360 (69.2) | 263 (66.4) | 345 (45.5) | 959 (67.2) | 771 (61.3) |
| **Southern U.S.** | 1,238 (32.7) | 933 (32.7) | 647 (35.9) | 186 (31.3) | 151 (27.4) | 158 (30.4) | 132 (33.3) | 410 (54.1) | 459 (32.1) | 475 (37.8) |
| **Born outside U.S.** | 11 (0.3) | 5 (0.2) | 7 (0.4) | 4 (0.7) | 1 (0.2) | 1 (0.2) | 1 (0.3) | 1 (0.1) | 1 (0.1) | 1 (0.1) |
| **U.S., region unspecified** | 6 (0.2) | 7 (0.2) | 5 (0.3) | 0 (0.0) | 0 (0.0) | 1 (0.2) | 0 (0.0) | 2 (0.3) | 9 (0.6) | 11 (0.9) |
| **Mother's education, years, mean (SD)** | 9.2 (3.3) | 8.9 (3.2) | 9.5 (3.4) | 9.6 (3.5) | 9.2 (3.3) | 9.3 (3.2) | 8.2 (3.9) | 7.7 (3.7) | 7.9 (3.1) | 7.7 (3.2) |
| **Missing, No. (%)** | 400 (10.5) | 311 (10.9) | 178 (9.8) | 42 (7.0) | 42 (7.6) | 52 (10.0) | 39 (9.8) | 122 (16.1) | 195 (13.6) | 194 (15.4) |
| **Father's education, years, mean (SD)** | 8.9 (3.5) | 8.6 (3.5) | 9.0 (3.7) | 9.0 (3.7) | 8.8 (3.7) | 8.9 (3.5) | 8.3 (4.1) | 7.7 (3.6) | 7.6 (3.3) | 7.5 (3.4) |
| **Missing, No. (%)** | 569 (15.0) | 406 (14.2) | 220 (12.2) | 74 (12.4) | 57 (10.3) | 56 (10.8) | 60 (15.2) | 164 (21.6) | 207 (14.5) | 225 (17.9) |
| **Participant education, years, mean (SD)** | 12.4 (3.0) | 12.0 (3.0) | 12.8 (3.1) | 12.7 (2.9) | 12.5 (2.8) | 12.7 (2.9) | 10.8 (3.1) | 10.2 (3.1) | 11.1 (3.3) | 10.7 (3.3) |
| **Childhood health, mean (SD)** | 1.8 (1.0) | 1.8 (1.0) | 1.8 (0.9) | 1.8 (1.0) | 1.8 (0.9) | 1.8 (1.0) | 1.9 (1.0) | 2.2 (1.2) | 1.9 (1.0) | 2.0 (1.0) |

**Note:** Transition-to-retirement clusters were defined as follows: (1) Retired after full-time work at ages 62–65; (2) Retired before age 60; (3) Retired after full-time work after age 70; (4) Full-time to part-time to retired after age 70; (5) Retired after part-time work at ages 62–65; (6) continuous part-time work; (7) mostly not working; (8) mostly disability; (9) unreported, then retired at ages 62–65; and (10) mostly unreported. Baseline characteristics are presented by transition-to-retirement cluster for participants included in the analytic sample. Age refers to participants' age at their first available dementia probability score during the outcome measurement period (age ≥76 years). Mother's and father's education were measured as years of schooling; missing indicators were included for participants with missing information on parental education. Childhood health was assessed retrospectively based on self-reported general health before age 16, ranging from 1 (excellent) to 5 (poor), with higher values indicating poorer childhood health. NH = non-Hispanic

### Association between transition-to-retirement clusters and dementia probability

The coefficients in Figure 3 show associations between transition-to-retirement clusters and dementia probability. The three clusters characterized by full-time work followed by retirement had similar odds of dementia: retirement after full-time work at ages 62–65 (the largest cluster and reference group), retirement before age 60 (OR: 1.03, 95% CI: 0.89, 1.20), and retirement after age 70 (OR: 1.05, 95% CI: 0.89, 1.24). Clusters characterized by part-time work generally had lower odds of dementia than the reference group. The “Full-time to part-time to retired after age 70” cluster (OR: 0.75, 95% CI: 0.58, 0.96) and the “Continuous part-time work” cluster (OR: 0.62, 95% CI: 0.48, 0.80) had significantly lower odds of dementia, while the “Retired after part-time work at ages 62–65” cluster had a similar but non-significant association (OR: 0.81, 95% CI: 0.62, 1.07). The “Mostly not working” cluster had similar odds of dementia to the reference group (OR: 1.08, 95% CI: 0.81, 1.43), while the “Mostly disability” cluster had higher odds (OR: 1.53, 95% CI: 1.22, 1.91). Among clusters characterized by periods of unreported employment information, the “Unreported, then retired at ages 62–65” cluster did not differ from the reference group (OR: 1.12, 95% CI: 0.94, 1.34), while the “Mostly unreported” cluster had higher odds of dementia (OR: 1.29, 95% CI: 1.03, 1.61). Results from sensitivity analyses using an alternative dissimilarity measure for the sequence analysis and alternative dementia probability scores were similar in direction and magnitude to those from the main analysis.

**Figure 3.**
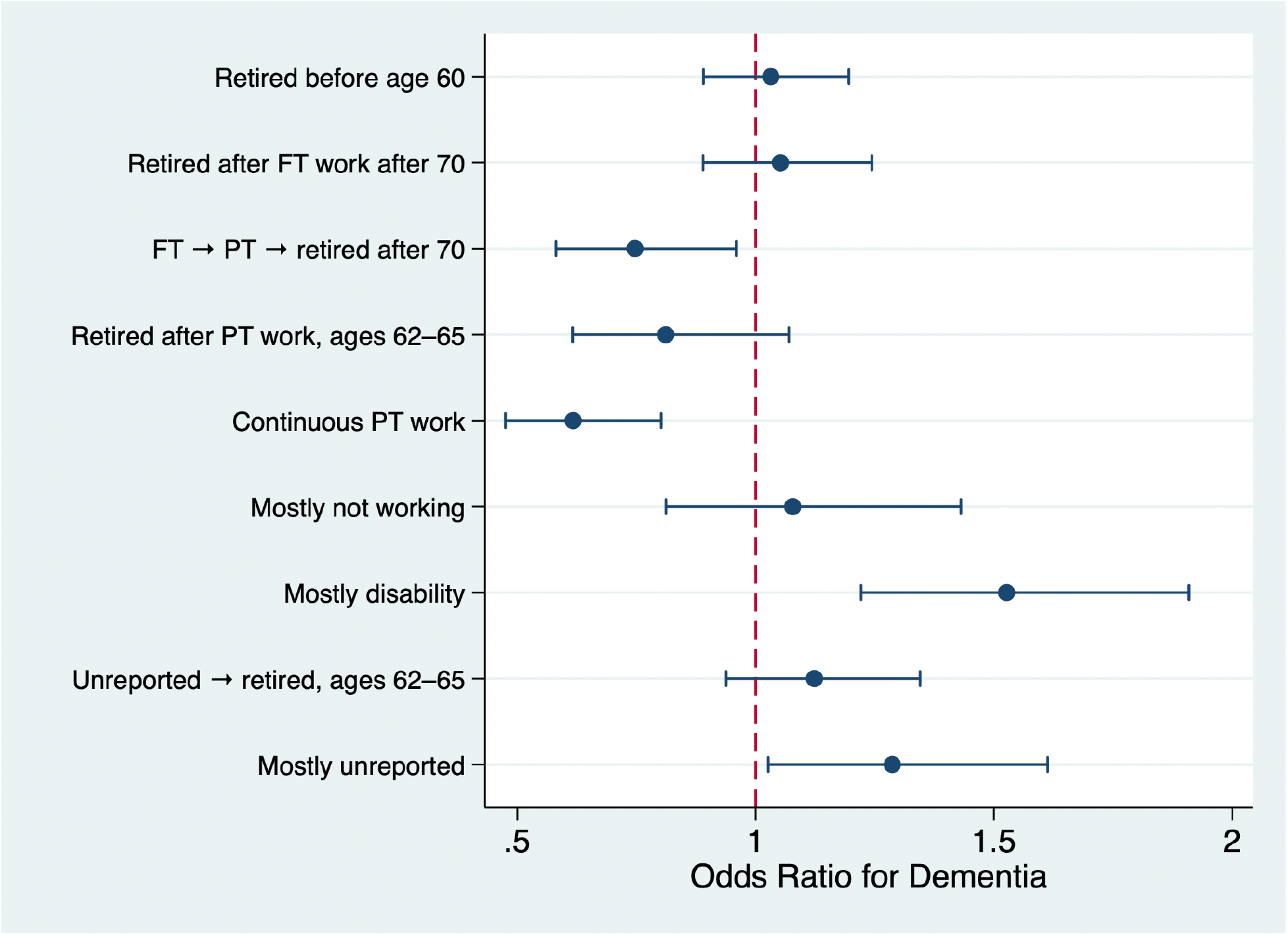
Associations between transition-to-retirement clusters and dementia probability. The coefficient plots show Odds Ratios and confidence intervals from a generalized estimating equations (GEE) analysis estimating the association between transition-to-retirement clusters and repeated dementia probability scores (ranging from 0-1). The model was adjusted for the following baseline confounders: age, gender, race and ethnicity, birth place, years of schooling, mother’s and father’s education, missing mother’s and father’s education, childhood health and survey wave. Of the 13,966 participants included in the sequence and cluster analysis, 87 were excluded due to missing covariates, which resulted in an analytic sample of 13,879 participants (49,360 person-waves). The model included inverse probability weights calculated at each time point.

## Discussion

Using data from a large diverse sample, we examined whether patterns of transition-to-retirement from ages 51-75 were associated with dementia risk at age 76 or older. Using sequence and cluster analysis, we were able to reduce thousands of individual transition-to-retirement trajectories into 10 clusters. We examined the timing of retirement and work intensity (full-time vs. part-time work) during the transition to retirement years to evaluate if they are differentially associated with dementia risk. We found that, compared with participants who transitioned directly from full-time work to retirement between ages 62 and 65, those who made the same transition either earlier or later had similar dementia risk. In contrast, transitioning from full-time to part-time work before retirement and continuous part-time between ages 51-75 were associated with lower dementia risk, while prolonged disability and prolonged periods of unreported employment information were associated with higher dementia risk.

Our findings on early retirement do not support the “mental retirement”^7^ or “use it or lose it”^6^ hypotheses and contrast with prior studies reporting worse cognitive outcomes among individuals who retire earlier.^7,9,29^ One key distinction from earlier work is our focus on complete retirement trajectories and long-term cognitive outcomes, rather than cognitive changes immediately following retirement. One possible explanation is that the effects of retirement on cognitive function attenuate following the initial transition period into retirement. Supporting this interpretation, an analysis using the Health and Retirement Study followed individuals from ages 51 to 75 and compared earlier versus later retirement showing substantial differences in cognitive functioning in the years immediately after retirement, but with differences becoming much smaller by 10 years post-retirement.^8^ Alternatively, associations between earlier retirement and poorer cognitive outcomes reported in prior studies may partly reflect health selection: individuals in poorer health may be more likely to retire earlier, while those who remain employed until older ages may be in better health and therefore at lower subsequent risk of dementia.

Our results also suggest that associations between later retirement and dementia risk may depend on work intensity. Findings on part-time work are novel and indicate that individuals who transitioned from full-time to part-time work before retirement and those consistently employed part-time had lower dementia risk than those who retired from full-time work between ages 62 and 65. These findings are consistent with life-course and cognitive reserve perspectives, suggesting that part-time work in later life may provide a balance between continued cognitive and social engagement and reduced occupational demands. Compared with full retirement, part-time work may help maintain cognitive stimulation, social interaction, and daily structure, while compared with full-time work, it may provide greater time for rest, leisure, and other health-promoting activities and reduce exposure to occupational stress.^10,30^ This interpretation is supported by our finding that later retirement after continuous full-time work was not associated with lower dementia risk. Alternatively, the association with part-time work may reflect health selection, as individuals who are able to continue working part-time until older ages may already be in better health and have higher cognitive functioning than those who fully retire.

The comparison between our findings on part-time work and prior studies using different time periods underscores the importance of the timing of part-time work for cognitive outcomes.^3,31,32^ A recent analysis using a subsample of the Health and Retirement Study found that a similar duration of part-time work earlier in life (between ages 30 and 60) was associated with a higher risk of dementia among both men and women.^3^ The divergence between these findings points to the role of contextual and structural features of employment across the lifecourse. Part-time work in early and mid-adulthood is more likely to reflect labor market disadvantages, including financial strain, job instability, and limited access to cognitively enriching work environments - factors that may constrain cognitive benefits.^33,34^ In later life, however, part-time work may serve as a bridge between full-time employment and retirement, potentially providing greater flexibility while maintaining social and cognitive engagement. At the same time, later-life part-time work is heterogeneous and may also reflect financial necessity, health limitations, or other constraints rather than a voluntary reduction in work intensity^35,36^. Future research should examine the mechanisms underlying the association between later-life part-time work and cognitive health, including the characteristics and cognitive demands of paid work, reasons for part-time employment, financial circumstances, and opportunities for social engagement and other cognitively stimulating activities outside of work.

Finally, the association between prolonged periods of disability during transition-to-retirement years is consistent with prior studies. Several mechanisms may contribute to this association. Disability in later life often results from chronic health conditions and sensory or functional impairments that may themselves be associated with dementia risk, including cardiovascular and neurological conditions, hearing and vision loss, and chronic pain^2,37–39^. Disability and mobility limitations may also reduce opportunities for physical activity and exercise, as well as social participation and engagement in cognitively stimulating activities, all of which may contribute to poorer cognitive health^40,41^. In addition, prolonged disability may reduce labor force participation and contribute to socioeconomic disadvantage, with potential downstream consequences for cognitive health^42,43^. Thus, the association observed in our study may reflect both the underlying health conditions leading to disability and the behavioral, social, and economic consequences of prolonged disability.

Our study has several strengths and offers important novel contributions to the current literature on cognitive function and work to retirement pathways. It uses sequence analysis, a novel approach for this area of research that allows for identifying meaningful differences between trajectories while at the same time grouping similar trajectories. Compared to other approaches for longitudinal data analysis, sequence analysis allows capturing differences across three different dimensions (timing, duration and order of events). It uses prospective information reported at each wave to define transition-to-retirement trajectories, which, compared to retrospective information, is less likely to be mis-reported.

Our study also has some limitations that should be considered when interpreting our study findings. First, because the trajectories included a combination of prospective and retrospective information, we could not include relevant confounders reported at the first wave such as income, marital status and type of work, which are likely to be associated with both retirement paths and cognition. Second, we had one cluster characterized by mostly unreported employment status, whose results were difficult to interpret. Third, the relationship between transition-to-retirement and cognition might be bi-directional, since individuals experiencing signs of dementia or cognitive impairment could be more likely to retire earlier. Fourth, because dementia probability was assessed starting at age 76, our analytic sample was restricted to individuals who survived and remained in the sample until that age. This may have introduced survivor selection, particularly if individuals who left employment early because of poor health or declining cognition were also more likely to die before reaching the outcome assessment period. Consequently, some employment trajectories characterized by early labor force exit due to poor health and premature mortality may be underrepresented in our analytic sample, potentially biasing associations between these retirement pathways and dementia risk.

## Conclusion

Using a large sample of U.S. older adults and sequence analysis, we found limited evidence of differential associations between retirement timing and dementia risk. In contrast, gradual retirement and continuous part-time employment between ages 51 and 75 were associated with lower dementia risk compared to retirement following full-time work. These associations should not be interpreted as causal. In particular, selection into retirement pathways and reverse causation may contribute to the observed associations: individuals with higher cognitive functioning, greater educational or occupational resources, or fewer health limitations may be both more able to remain employed part-time or transition gradually into retirement and less likely to subsequently develop dementia. Future studies should distinguish these selection processes from potential benefits of gradual retirement and identify whether continued cognitive and social engagement, occupational demands, work flexibility, and opportunities for other health-promoting activities contribute to cognitive health in later life.

In contrast, individuals with extended periods of disability during the transition-to-retirement years had higher dementia risk than those retiring from full-time work. Identifying the health, social, and economic mechanisms underlying this association may help inform strategies to support cognitive health among older adults experiencing prolonged disability. Future research is needed to assess the robustness of these findings across contexts and isolate specific underlying mechanisms.

## Ethics Statement

This study used publicly available, anonymized data from the Health and Retirement Study (HRS). HRS participants provided informed consent for participation in the original study. The present study involved secondary analysis of existing anonymized data and did not include identifiable personal information. Therefore, ethical approval was not required for this study.

## Data availability statement

Data used in this study are available from the Health and Retirement Study (HRS). Public-use HRS data are available to registered users through the HRS website. Access to restricted data, where applicable, requires application and approval by the HRS.

## Funding

This research was supported by the National Institute of Health/National Institute on Aging (award: R01AG074351, PI Vable)

## Notes

### Competing Interest Statement

The authors have declared no competing interest.

### Author Declarations

The study used ONLY openly available human data that are available through the Health and Retirement Study.

## References

1. Fang, M. et al. Lifetime risk and projected burden of dementia. Nat Med 31, 772–776 (2025).

2. Livingston, G. et al. Dementia prevention, intervention, and care: 2024 report of the Lancet standing Commission. The Lancet 404, 572–628 (2024).

3. Pacca, L. et al. Do type, timing and duration of life course non-employment differentially predict dementia risk? An application of sequence analysis. Social Science & Medicine 372, 117976 (2025).

4. Huang, L.-Y. et al. Association of Occupational Factors and Dementia or Cognitive Impairment: A Systematic Review and Meta-Analysis. JAD 78, 217–227 (2020).

5. Transamerica Center for Retirement Studies. Retirement in the USA: The Outlook of the Workforce. https://www.transamericainstitute.org/research/publications/details/retirement-in-the-usa-workforce-outlook-survey-report-2025 (2025).

6. Hultsch, D. F., Hertzog, C., Small, B. J. & Dixon, R. A. Use it or lose it: Engaged lifestyle as a buffer of cognitive decline in aging? Psychology and Aging 14, 245–263 (1999).

7. Rohwedder, S. & Willis, R. J. Mental Retirement. Journal of Economic Perspectives 24, 119–138 (2010).

8. Bonsang, E., Adam, S. & Perelman, S. Does retirement affect cognitive functioning? Journal of Health Economics 31, 490–501 (2012).

9. Mazzonna, F. & Peracchi, F. Unhealthy Retirement? J. Human Resources 52, 128–151 (2017).

10. Baumann, I. et al. Do cognitively stimulating activities affect the association between retirement timing and cognitive functioning in old age? Ageing and Society 42, 306–330 (2022).

11. Yauk, J., Veal, B. & Dobbs, D. Understanding the Link Between Retirement Timing and Cognition: A Scoping Review. J Appl Gerontol 43, 588–600 (2024).

12. Celidoni, M., Dal Bianco, C. & Weber, G. Retirement and cognitive decline. A longitudinal analysis using SHARE data. Journal of Health Economics 56, 113–125 (2017).

13. Clouston, S. A. P. & Denier, N. Mental retirement and health selection: Analyses from the U.S. Health and Retirement Study. Social Science & Medicine 178, 78–86 (2017).

14. Denier, N., Clouston, S. A. P., Richards, M. & Hofer, S. M. Retirement and cognition: A life course view. Advances in Life Course Research 31, 11–21 (2017).

15. Carr, D. C., Matz, C., Taylor, M. G. & Gonzales, E. Retirement Transitions in the United States: Patterns and Pathways From Full-Time Work. Public Policy & Aging Report 31, 71–77 (2021).

16. Maestas, N. Back to Work: Expectations and Realizations of Work after Retirement. J. Human Resources 45, 718–748 (2010).

17. Juster, R.-P., McEwen, B. S. & Lupien, S. J. Allostatic load biomarkers of chronic stress and impact on health and cognition. Neuroscience & Biobehavioral Reviews 35, 2–16 (2010).

18. Carr, D. C., Willis, R., Kail, B. L. & Carstensen, L. L. Alternative Retirement Paths and Cognitive Performance: Exploring the Role of Preretirement Job Complexity. The Gerontologist 60, 460–471 (2020).

19. Hoven, H., Dragano, N., Blane, D. & Wahrendorf, M. Early Adversity and Late Life Employment History—A Sequence Analysis Based on SHARE. Work, Aging and Retirement 4, 238–250 (2018).

20. Wahrendorf, M., Zaninotto, P., Hoven, H., Head, J. & Carr, E. Late Life Employment Histories and Their Association With Work and Family Formation During Adulthood: A Sequence Analysis Based on ELSA. The Journals of Gerontology: Series B 73, 1263–1277 (2018).

21. Calvo, E., Madero-Cabib, I. & Staudinger, U. M. Retirement Sequences of Older Americans: Moderately Destandardized and Highly Stratified Across Gender, Class, and Race. The Gerontologist 58, 1166–1176 (2018).

22. Pacca, L. et al. Using sequence and cluster analysis to characterize variables that unfold over time: implementation and practical considerations for epidemiologists. American Journal of Epidemiology 195, 1707–1718 (2026).

23. Abbott, A. & Forrest, J. Optimal Matching Methods for Historical Sequences. Journal of Interdisciplinary History 16, 471 (1986).

24. Gianattasio, K. Z., Ciarleglio, A. & Power, M. C. Development of Algorithmic Dementia Ascertainment for Racial/Ethnic Disparities Research in the US Health and Retirement Study. Epidemiology 31, 126–133 (2020).

25. Vable, A. M., Gilsanz, P., Nguyen, T. T., Kawachi, I. & Glymour, M. M. Validation of a theoretically motivated approach to measuring childhood socioeconomic circumstances in the Health and Retirement Study. PLoS ONE 12, e0185898 (2017).

26. Daza, E. J., Hudgens, M. G. & Herring, A. H. Estimating Inverse-probability Weights for Longitudinal Data with Dropout or Truncation: The Xtrccipw Command. The Stata Journal: Promoting communications on statistics and Stata 17, 253–278 (2017).

27. Hamming, R. W. Error Detecting and Error Correcting Codes. Bell System Technical Journal 29, 147–160 (1950).

28. Studer, M. & Ritschard, G. What Matters in Differences Between Life Trajectories: A Comparative Review of Sequence Dissimilarity Measures. Journal of the Royal Statistical Society Series A: Statistics in Society 179, 481–511 (2016).

29. Mazzonna, F. & Peracchi, F. Ageing, cognitive abilities and retirement. European Economic Review 56, 691–710 (2012).

30. Van Solinge, H., Vanajan, A. & Henkens, K. Does Phased Retirement Increase Vitality in Older Workers? Findings from a 3-Year Follow-Up Panel Study. Journal of Aging & Social Policy 35, 221–240 (2023).

31. Ice, E., Ang, S., Greenberg, K. & Burgard, S. Women’s Work-Family Histories and Cognitive Performance in Later Life. American Journal of Epidemiology 189, 922–930 (2020).

32. Bertogg, A. & Leist, A. K. Gendered life courses and cognitive functioning in later life: the role of context-specific gender norms and lifetime employment. Eur J Ageing 20, 7 (2023).

33. Gallie, D. et al. Quality of work and job satisfaction: comparing female part-time work in four European countries. International Review of Sociology 26, 457–481 (2016).

34. OECD. OECD Employment Outlook 2020: Worker Security and the COVID-19 Crisis. (OECD Publishing, 2020). doi:10.1787/1686c758-en.

35. Dingemans, E. & Henkens, K. Involuntary retirement, bridge employment, and satisfaction with life: A longitudinal investigation: RETIREMENT TRANSITIONS AND LIFE SATISFACTION. J. Organiz. Behav. 35, 575–591 (2014).

36. Cahill, K., Giandrea, M., Quinn, J., Sacco, L. & Platts, L. Does Bridge Employment Mitigate or Exacerbate Inequalities Later in Life? Innovation in Aging 5, 414–414 (2021).

37. Loughrey, D. G., Kelly, M. E., Kelley, G. A., Brennan, S. & Lawlor, B. A. Association of Age-Related Hearing Loss With Cognitive Function, Cognitive Impairment, and Dementia: A Systematic Review and Meta-analysis. JAMA Otolaryngol Head Neck Surg 144, 115 (2018).

38. Shang, X., Zhu, Z., Wang, W., Ha, J. & He, M. The Association between Vision Impairment and Incidence of Dementia and Cognitive Impairment. Ophthalmology 128, 1135–1149 (2021).

39. Tian, J. et al. Association between chronic pain and risk of incident dementia: findings from a prospective cohort. BMC Med 21, 169 (2023).

40. Cao, B.-F. et al. Association Between Mobility Limitations and Cognitive Decline in Community-Dwelling Older Adults: The English Longitudinal Study of Ageing. The Gerontologist 64, gnae139 (2024).

41. Grande, G., Qiu, C. & Fratiglioni, L. Prevention of dementia in an ageing world: Evidence and biological rationale. Ageing Research Reviews 64, 101045 (2020).

42. Bodryzlova, Y. et al. Social class and the risk of dementia: A systematic review and meta-analysis of the prospective longitudinal studies. Scand J Public Health 51, 1122–1135 (2023).

43. Wang, M., Zhan, Y., Liu, S. & Shultz, K. S. Antecedents of bridge employment: A longitudinal investigation. Journal of Applied Psychology 93, 818–830 (2008).

