## Supplemental Material for "The transition to retirement and dementia risk: An application of sequence analysis"

### Figure A1 - Study sample selection criteria

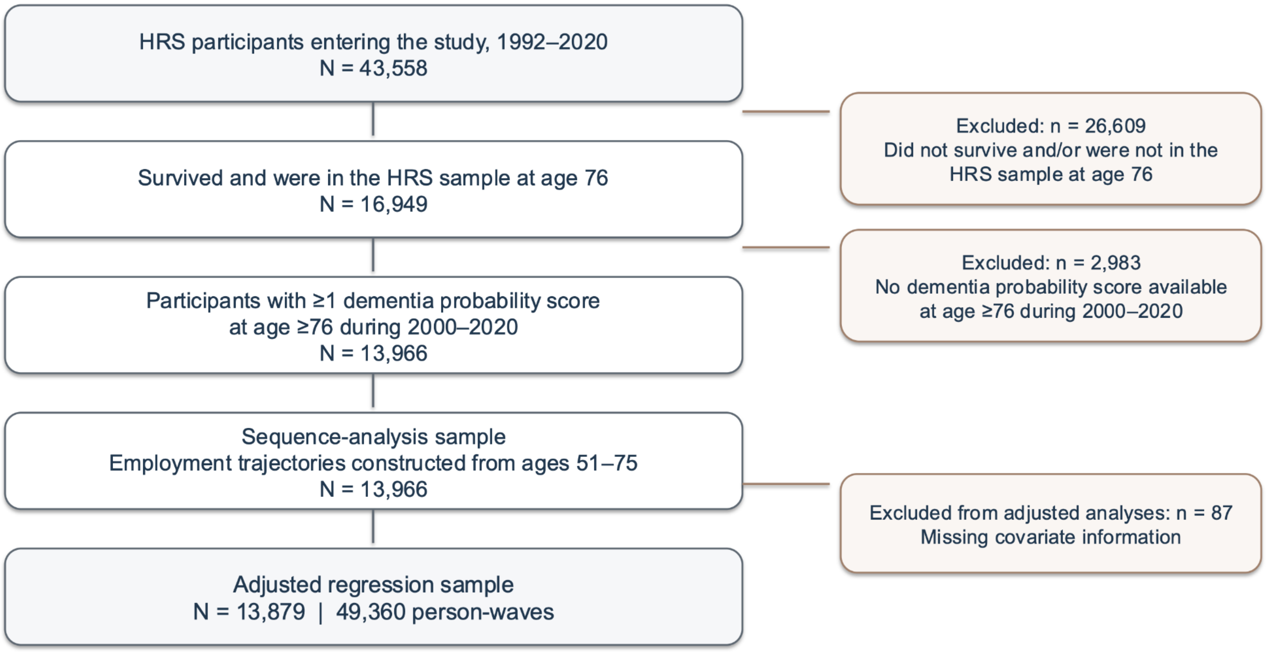

**Figure A1 - Cluster quality indicators using the Optimal Matching dissimilarity measure (main analysis)**

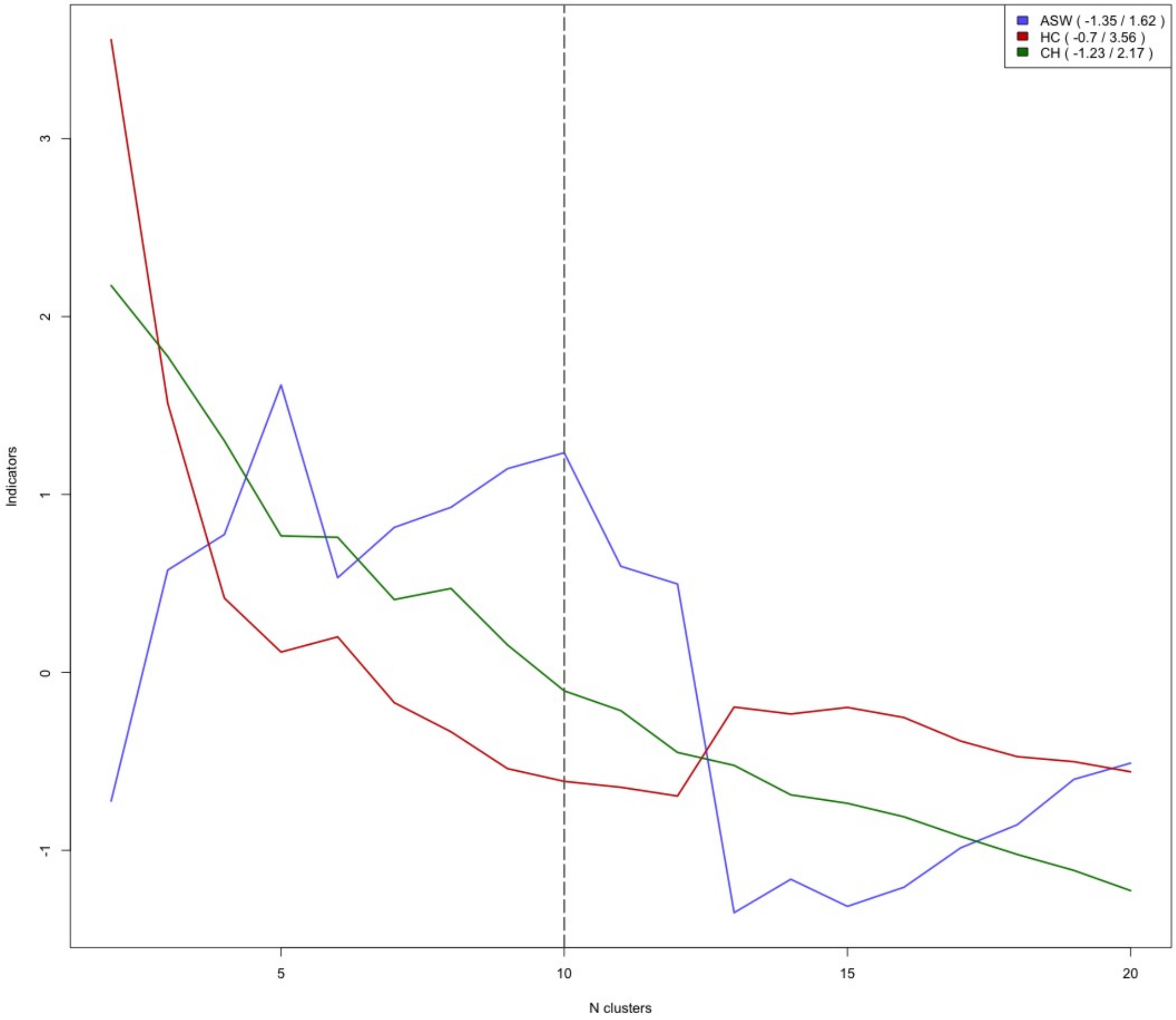


This figure graphically visualizes three commonly used cluster quality measures (Average Silhouette Width, ASW, Hubert & Levin C index, HC and Calinski-Harabasz index, CH) for a chosen range of partitions (2-20 clusters) obtained using hierarchical clustering algorithm to group transition-to-retirement trajectories after conducting sequence analysis using the Optimal Matching algorithm. All three measures were z-standardized to improve the plot's readability. For better cluster quality, the goal is to maximize the ASW and CH and minimize HC. Based on this guideline, we selected 10 clusters as our main cluster solution.

**Figure A2 – Cluster quality indicators using the Hamming dissimilarity measure (sensitivity analysis)**

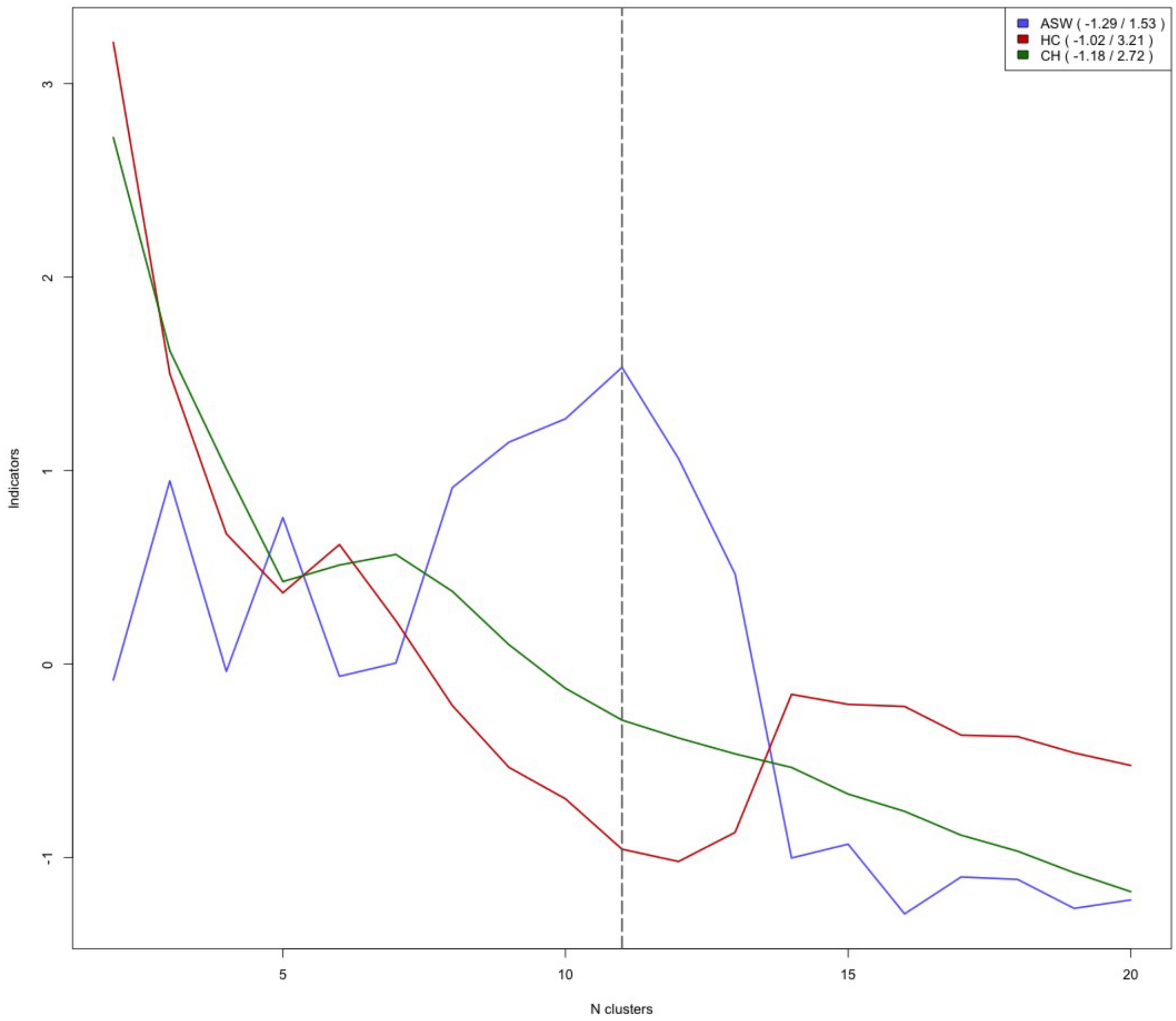


This figure graphically visualizes three commonly used cluster quality measures (Average Silhouette Width, ASW, Hubert & Levin C index, HC and Calinski-Harabasz index, CH) for a chosen range of partitions (2-20 clusters) obtained using hierarchical clustering algorithm to group transition-to-retirement trajectories after conducting sequence analysis using the Hamming algorithm. All three measures were z-standardized to improve the plot's readability. For better cluster quality, the goal is to maximize the ASW and CH and minimize HC. Based on this guideline, we selected 11 clusters for our sensitivity analysis.

**Figure A3 - Sequence index plots by cluster using the Hamming algorithm (sensitivity analysis)**

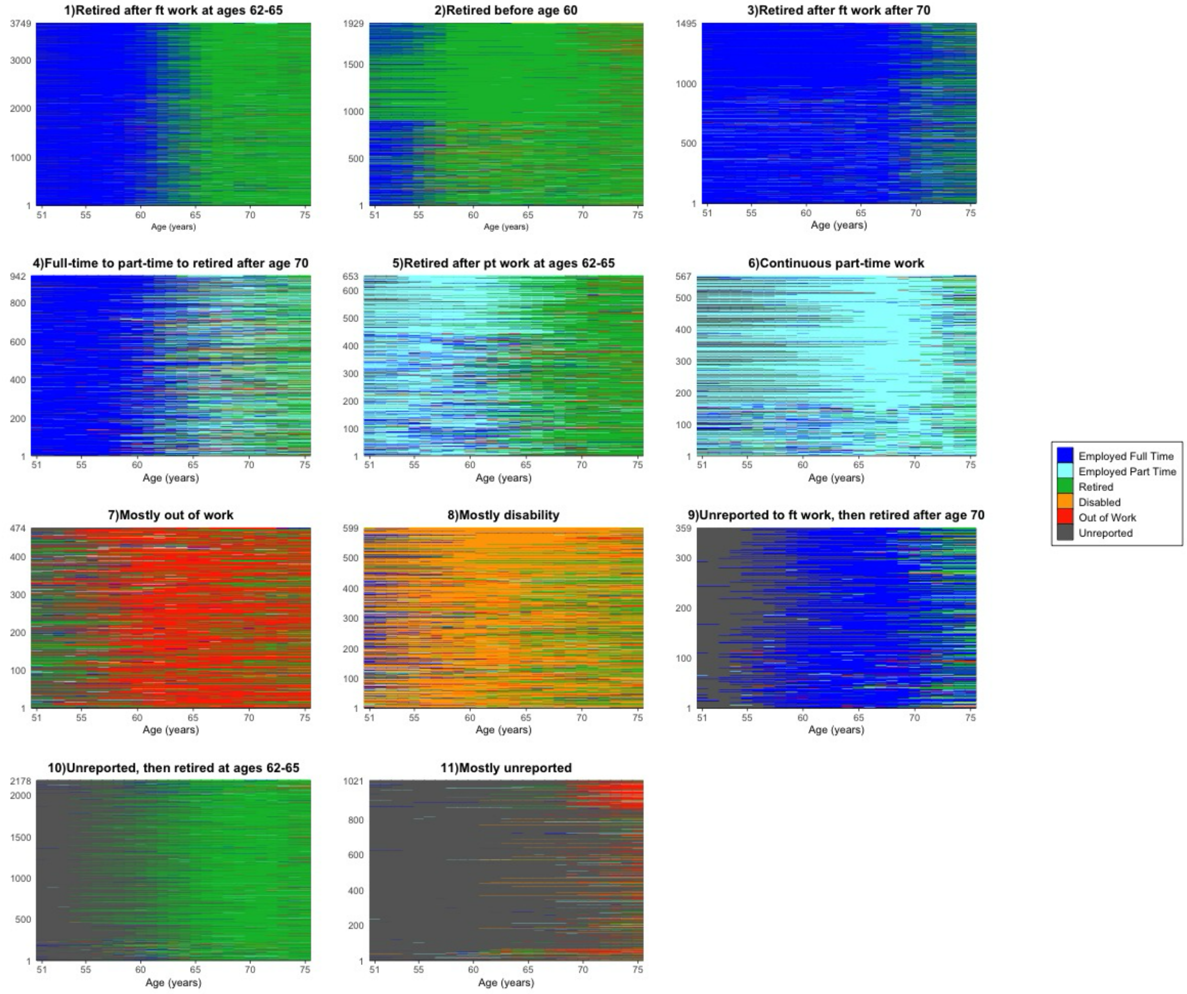


Sequence index plots for the transition-to-retirement clusters obtained using sequence and cluster analysis among HRS participants who had at least one wave of valid dementia probability score at age 76 or older (N=13,966). We applied the Hamming algorithm to calculate the dissimilarity between sequences, using substitution costs based on transition frequencies. We then implemented hierarchical clustering to group sequences based on their similarity. Based on cluster quality indicators, we identified 10 transition-to-retirement clusters. Each index plot shows the number of participants on the y-axis and age (51-75) on the x-axis.

**Figure A4 – Association between transition-to-retirement clusters obtained using the Hamming algorithm and dementia probability (sensitivity analysis)**

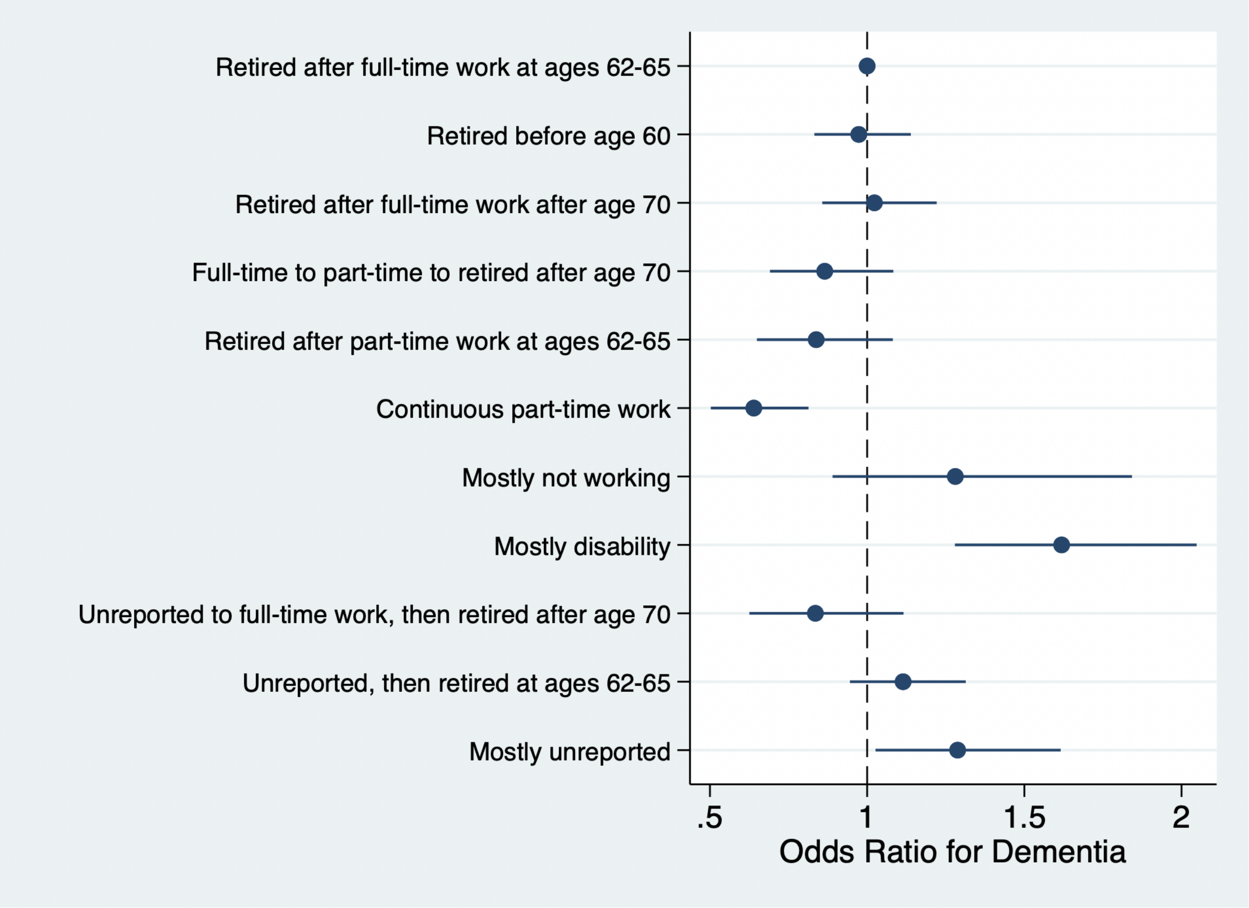


The coefficient plots show Odds Ratios and confidence intervals from a generalized estimating equations (GEE) analysis estimating the association between transition-to-retirement clusters obtained using the Hamming algorithm (sensitivity analysis) and repeated dementia probability scores (ranging from 0-1). The model was adjusted for the following baseline confounders: age, gender, race and ethnicity, birth place, years of schooling, mother's and father's education, missing mother's and father's education, childhood health and survey wave. Of the 13,966 participants included in the sequence and cluster analysis, 87 were excluded due to missing covariates, which resulted in an analytic sample of 13,879 participants (49,360 person-waves). The model included inverse probability weights calculated at each time point.

**Figure A5 – Association between transition-to-retirement clusters and Hudomiet dementia probability scores (sensitivity analysis)**

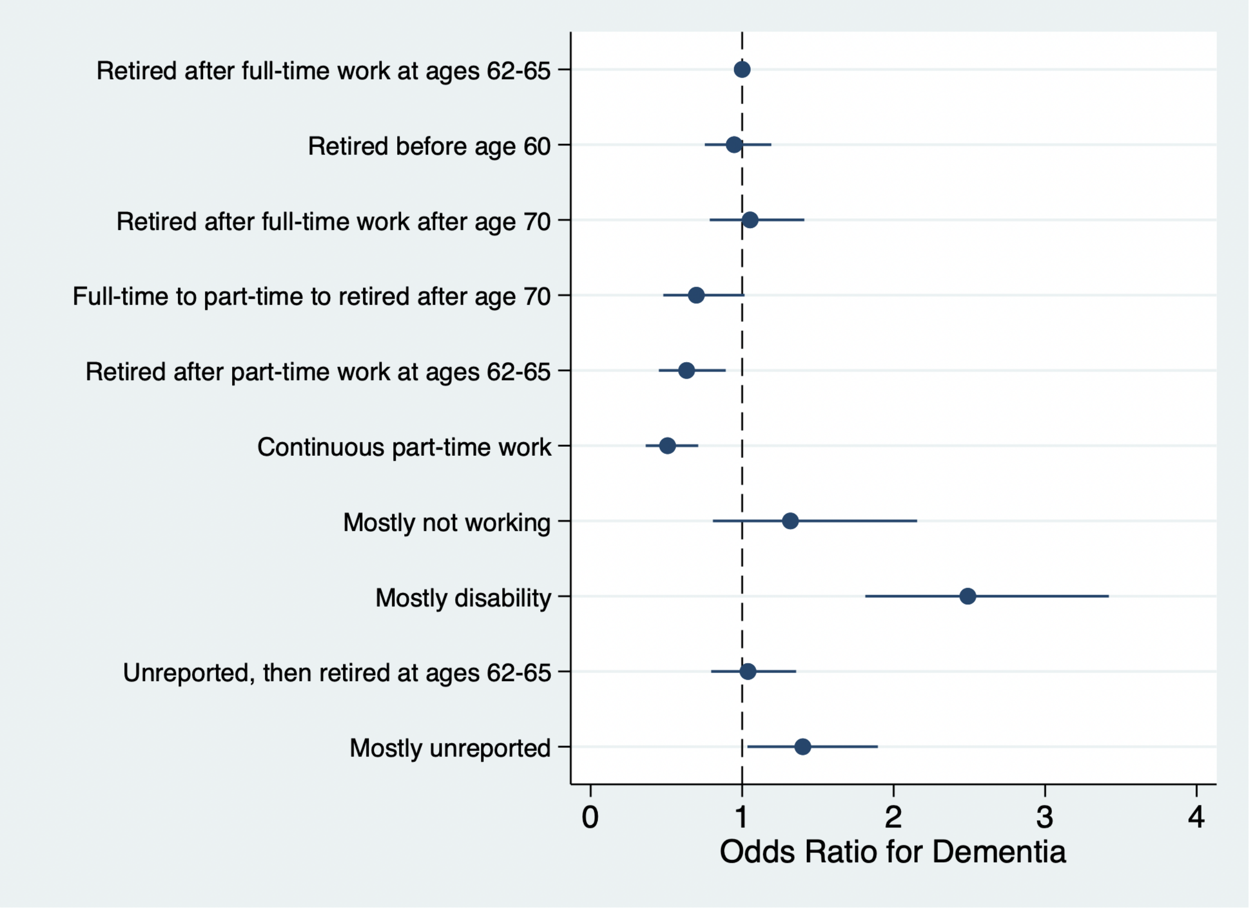


The coefficient plots show Odds Ratios and confidence intervals from a generalized estimating equations (GEE) analysis estimating the association between transition-to-retirement clusters and repeated dementia probability scores (ranging from 0-1) developed by Hudomiet and colleagues. The model was adjusted for the following baseline confounders: age, gender, race and ethnicity, birthplace, years of schooling, mother's and father's education, missing mother's and father's education, childhood health and survey wave. This sensitivity analysis was conducted using an analytic sample of 12,567 participants (42,640 person-waves). Since this measure is only available until 2016 (vs. 2020 for the Gianattasio measure), the analytic sample was smaller than in the main analysis. The model included inverse probability weights calculated at each time point.
